# Towards Maps of Disease Progression: Biomedical Large Language Model Latent Spaces For Representing Disease Phenotypes And Pseudotime

**DOI:** 10.1101/2024.06.16.24308979

**Authors:** Rafael Zamora-Resendiz, Ifrah Khurram, Silvia Crivelli

## Abstract

In this study, we propose a scientific framework to detect capability among biomedical large language models (LLMs) for organizing expressions of comorbid disease and temporal progression. We hypothesize that biomedical LLMs pretrained on next-token prediction produce latent spaces that implicitly capture "disease states" and disease progression, i.e., the transitions over disease states over time. We describe how foundation models may capture and transfer knowledge from explicit pretraining tasks to specific clinical applications. A scoring function based on Kullback-Leibler divergence was developed to measure "surprise" in seeing specialization when subsetting admissions along 13 biomedical LLM latent spaces. By detecting implicit ordering of longitudinal data, we aim to understand how these models self-organize clinical information and support tasks such as phenotypic classification and mortality prediction. We test our hypothesis along a case study for obstructive sleep apnea (OSA) in the publicly available MIMIC-IV dataset, finding ordering of phenotypic clusters and temporality within latent spaces. Our quantitative findings suggest that increased compute, conformance with compute-optimal training, and widening contexts promote better implicit ordering of clinical admissions by disease states, explaining 60.3% of the variance in our proposed implicit task. Preliminary qualitative findings suggest LLMs’ latent spaces trace patient trajectories through different phenotypic clusters, terminating at end-of-life phenotypes. This approach highlights the potential of biomedical LLMs in modeling disease progression, identifying new patterns in disease pathways and interventions, and evaluating clinical hypotheses related to drivers of severe illness. We underscore the need for larger, high-resolution longitudinal datasets to further validate and enhance understanding of the utility of LLMs in modeling patient trajectories along clinical text and advancing precision medicine.

**Key Points:** *Question:* Do LLMs sensibly organize cilnical data with respect to applications in precision medicine?

*Findings:* Biomedically-trained LLMs show increasing potential in promoting the organization of patient data to reflect disease progression. In a subcohort of OSA patients, maps derived from LLMs’ latent representations reveal traceable disease trajectories.

*Meaning:* Maps of disease progression offer an explanation to the utility of LLMs in precision medicine. Following current pretraining conventions in foundation modeling, scientific inquiry into these maps may help anticipate progress in applications of LLMs for healthcare.

## Introduction

Computational medicine has made important strides over the past two decades with the digitization and centralization of patient medical data into electronic health records (EHR)^1,2^. Even so, EHR systems are primarily designed, implemented, and operationalized for administrative function. Common structured medical variables including diagnosis codes, laboratory orders and results, procedure codes, and medication prescriptions typically focus on billable procedures. As a result, relevant patient phenotypes for predictive modeling are under-represented or poorly characterized in structured EHR. This leads to substantial work in order to harmonize clinical concepts and biomarkers using both structured EHR^3^ and unstructured text. A means of integrating health status information locked in clinical text with structured representations of medical knowledge is through the application of natural language processing (NLP)^2,4^. Recently in clinical medicine, large-language models (LLMs) have been adopted to process clinical text for topic categorization^5^, medical concept extraction^6,7^, and medical question-answering^8^.

While LLMs continue to show aptitude for general NLP applications as model size increase from millions to trillions of trainable parameters and pretraining scales to tens of terabytes of text^9^, clinically pretrained LLMs lag behind current state-of-the-art (SOTA) model pretraining on non-medical text. As the field continues to move towards even larger LLMs^10^ and infusion with other socio-economic and biomedical modalities including census tract-level data^11^, medical imaging^12^, and genetics^13^, there remain open questions about the representative capabilities of LLMs in capturing the unique nuances of patient health. In particular, challenges remain in identifying how latent spaces map to structured clinical variables like demographics, disease phenotypes, medical interventions, and measures of life expectancy. Additionally, there lacks understanding on the effect of model pretraining in building coherent representations of these common determinants of health even with continued work in interpretability^14^ and clinical decision support^15^ for medical machine learning (ML) systems.

With the increased accessibility of large-scale EHR for A.I. research^2^, there exists greater opportunity to observe the effects of LLM scaling in large population-wide studies. We hypothesize that LLMs pretrained on biomedical subject matter capture medically coherent representations of clinical text in their latent spaces, facilitating clinically-relevant subsetting of patient records when embedding population-wide longitudinal corpora. We propose a mixed-method approach to develop an explanatory framework on the effectiveness of contemporary biomedical LLMs in modeling clinical phenotypes and trajectories. A latent space analysis over the publicly available MIMIC-IV clinical dataset^16^ was performed across 13 biomedical LLMs of different model sizes and pre-training strategies including BioBERT, BioBART, ClinicalBert, RadBERT, BioMegatron, Gatortron, BioGPT, and Forge-bio^10,17–23^. The proposed framework first employs a quantitative assessment detecting organizaton in LLM latent spaces with respect to sex, age, race, diagnosis codes, procedure codes, and death statistics. This is proceeded by qualitative assessment of the LLM latent spaces to interpret the coherence between corpus subsets and structured medical features by visualizing admission-level corpus manifolds, identifying medically-relevant subsetting, and manually reviewing clinical discharge reports for evidence of longitudinal ordering among a case study for obstructive sleep apnea (OSA).

We demonstrate application of the framework for investigating mechanistic drivers of severe disease in a sub-cohort of patients with diagnosed OSA^24^. OSA, a chronic illness causing interrupted breathing during sleep due to over-relaxation of muscles around the airway, is linked to several illnesses. This study investigates 6 diseases comorbid with OSA. Previous studies hypothesize that obstruction during sleep causes coronary blood flow to increase disproportionately with myocardial work increasing coronary artery vascular resistance, which may predict increased cardiovascular comorbidities in a dose-dependent relation. Understanding the interaction of OSA with these comorbidities is crucial for determining the additive risk of OSA in developing severe illnesses^25^. The case study focuses on discharge reports exhibiting severe OSA and concurrent heart failure (HF), finding that admissions indexed by probability of HF diagnosis along LLM latent spaces increase the odds ratio of textual mentions referencing dyspnea symptoms and ongoing OSA treatment. Additionally, potential non-adherence to OSA treatment, such as continuous positive airway pressure (CPAP) treatment, was observed, highlighting the utility of using clinical text to construct surrogates for prolonged exposure of respiratory obstruction.

## Methods

### Data

The analysis utilized the publicly available MIMIC-IV Clinical Database^26^ which includes admissions to the intensive care units (ICUs) at Boston’s Beth Israel Deaconess Medical Center from 2001 to 2019. EHR for 331,794 admissions, each with a recorded discharge report, were gathered, covering 1.6 million patient-days for a population of 145,915 patients. The unstructured reports were linked to structured medical data including admission-level demographics, billable procedures, billable diagnoses, and mortality outcomes (MIMIC-IV censors time of death past one year after the last admission). The distribution of patient-level demographics is reported in Table 1.

**Table 1.** Cohort Characteristics For MIMIC-IV Admissions with Discharge Report.

|  |  | Overall (n=145,915) | Non-OSA (n=131,588) | OSA (n=14,327) |
| --- | --- | --- | --- | --- |
| Sex | Male | 71,016 (48.7%) | 62,469 (47.5%) | 8,547 (59.7%) |
|  | Female | 74,899 (51.3%) | 69,119 (52.5%) | 5,780 (40.3%) |
| Race | White | 100,085 (68.6%) | 89,898 (68.3%) | 10,187 (71.1%) |
|  | Black | 17,475 (12.0%) | 15,456 (11.7%) | 2,019 (14.1%) |
|  | Other | 20,929 (14.3%) | 19,317 (14.7%) | 1,612 (11.3%) |
|  | Unknown | 7,426 (5.1%) | 6,917 (5.3%) | 509 (3.6%) |
| Hispanic Ethnicity | Yes | 7,006 (4.8%) | 6,322 (4.8%) | 684 (4.8%) |
|  | No | 131,483 (90.1%) | 118,349 (89.9%) | 13,134 (91.7%) |
|  | Unknown | 7,426 (5.1%) | 6,917 (5.3%) | 509 (3.6%) |
| Age at Death | All | 27,247 (18.7%) | 24,700 (18.8%) | 2,547 (17.8%) |
|  | < 20 | 9 (0.0%) | 9 (0.0%) | 0 (0.0%) |
|  | 20-29 | 215 (0.1%) | 204 (0.2%) | 11 (0.1%) |
|  | 30-39 | 415 (0.3%) | 379 (0.3%) | 36 (0.3%) |
|  | 40-49 | 1,090 (0.7%) | 995 (0.8%) | 95 (0.7%) |
|  | 50-59 | 2,960 (2.0%) | 2,654 (2.0%) | 306 (2.1%) |
|  | 60-69 | 5,014 (3.4%) | 4,343 (3.3%) | 671 (4.7%) |
|  | 70-79 | 6,128 (4.2%) | 5,375 (4.1%) | 753 (5.3%) |
|  | 80-89 | 7,329 (5.0%) | 6,787 (5.2%) | 542 (3.8%) |
|  | > 89 | 4,087 (2.8%) | 3,954 (3.0%) | 133 (0.9%) |
| Diagnosis | Stroke (STRK) | 18,719 (12.8%) | 16,471 (12.5%) | 2,248 (15.7%) |
|  | Heart Failure (HF) | 23,835 (16.3%) | 19,519 (14.8%) | 4,316 (30.1%) |
|  | Coronary Artery Disease (CAD) | 35,134 (24.1%) | 29,891 (22.7%) | 5,243 (36.6%) |
|  | Atrial Fibrillation (AFIB) | 26,048 (17.9%) | 22,179 (16.9%) | 3,869 (27.0%) |
|  | Type-2 Diabetes Mellitus (T2DB) | 32,752 (22.4%) | 26,730 (20.3%) | 6,022 (42.0%) |
|  | Hypertension (HTN) | 81,640 (55.9%) | 70,516 (53.6%) | 11,124 (77.6%) |
| Text Corpus (MB) |  | 3,338 | 2,781 | 557 |
| Admitted Patient-Days |  | 1,630,924 | 1,372,168 | 258,756 |
| Admissions | All | 331,794 | 280,229 | 51,565 |
|  | Emergency | 291,255 (87.8%) | 247,634 (88.4%) | 43,621 (84.6%) |
|  | Elective | 40,539 (12.2%) | 32,595 (11.6%) | 7,944 (15.4%) |
| Admissions By Age | < 20 | 1,410 (0.4%) | 1,402 (0.5%) | 8 (0.0%) |
|  | 20-29 | 18,821 (5.7%) | 18,115 (6.5%) | 706 (1.4%) |
|  | 30-39 | 25,170 (7.6%) | 22,999 (8.2%) | 2,171 (4.2%) |
|  | 40-49 | 35,634 (10.7%) | 30,223 (10.8%) | 5,411 (10.5%) |
|  | 50-59 | 59,674 (18.0%) | 47,971 (17.1%) | 11,703 (22.7%) |
|  | 60-69 | 70,549 (21.3%) | 55,336 (19.7%) | 15,213 (29.5%) |
|  | 70-79 | 59,461 (17.9%) | 48,681 (17.4%) | 10,780 (20.9%) |
|  | 80-89 | 45,347 (13.7%) | 40,556 (14.5%) | 4,791 (9.3%) |
|  | > 89 | 15,728 (4.7%) | 14,946 (5.3%) | 782 (1.5%) |

It is important to note that MIMIC-IV contains additional sources of unstructured data, such as radiology reports, but these were not considered in this analysis. The complete corpus of discharge reports totals 3.3 GB of text. Phenotypes for admissions were labeled for the presence of OSA, HF, stroke (STRK), atrial fibrillation (AFIB), coronary artery disease (CAD), type 2 diabetes mellitus (T2DB), and hypertension (HTN) according to the International Classification of Diseases, 9th and 10th Revision (ICD-9/10). Expert-curated ICD definitions for OSA and comorbid diagnoses are found in Table 2, and the rates of these conditions in the MIMIC-IV patient population are also reported in Table 1.

**Table 2.**
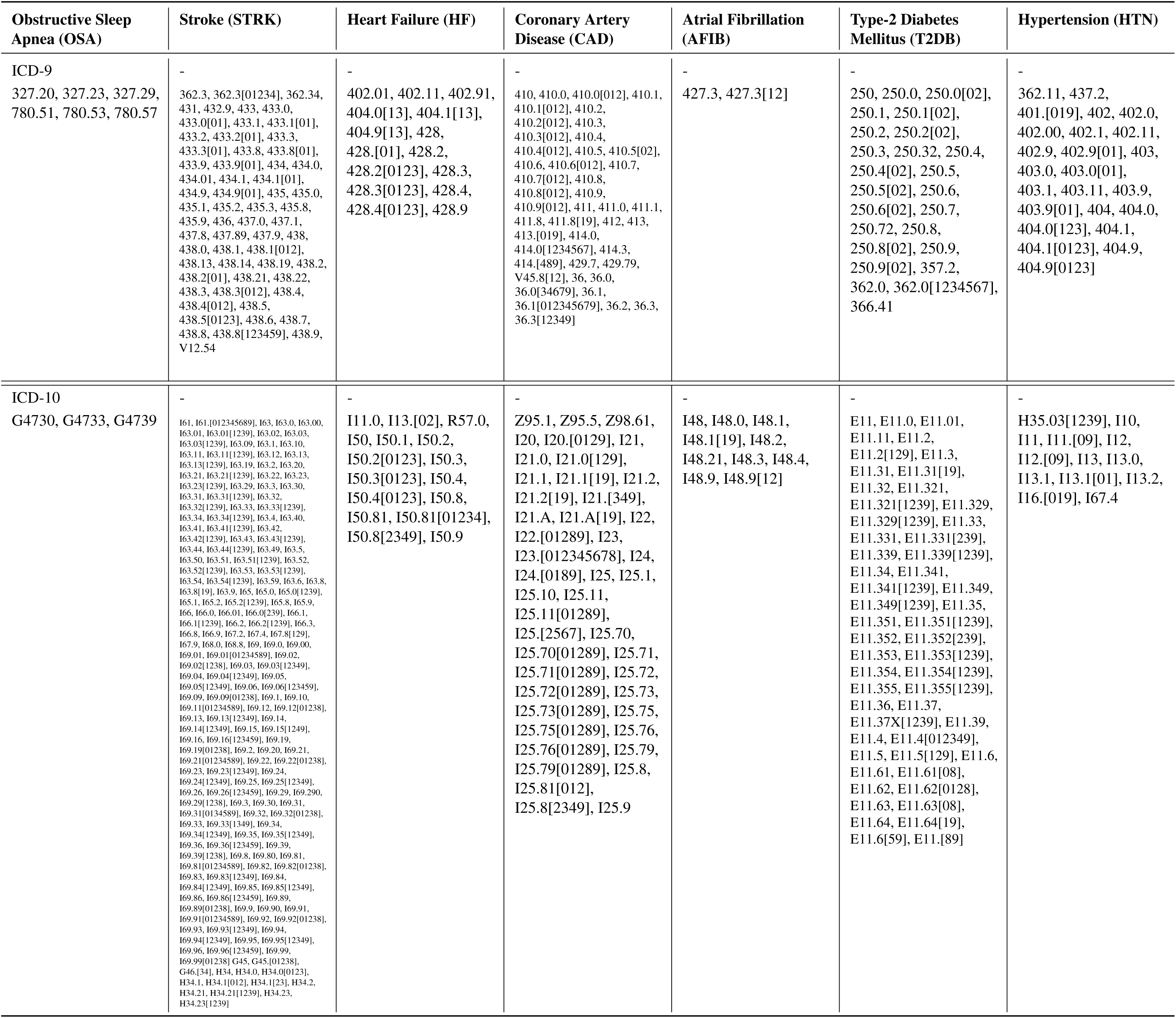
ICD 9/10 Definitions For OSA and Comorbid Diseases.

### Large Language Models

LLMs are attention-based deep learning architectures designed to predict the next token or missing tokens in sequentially ordered data^27^. An advantage of transformer-based language models over recurrent architectures like RNNs^28^ is the absence of recursive operations for computing next-token probability scores. Instead, transformers utilize parallelizable feed-forward operations. The computational graph for LLM implementations can be naively summarized as stacked blocks of multi-head self-attention followed by feed-forward multilayer perceptrons (MLP) with residual connections between inputs and outputs for both operations.

In essence, multi-head self-attention aggregates information from different sections of a sequence and the MLP projects this aggregated information into a new feature space. Adding more blocks or increasing the number of parameters per block enhances the language model’s ability to condition next-token prediction. However, scaling these models requires substantial computational resources and a variety of optimization techniques to manage numerical instability during pretraining^29^. Models of this size heavily rely on data-parallel, model-parallel, and pipeline-parallel training, given that a single GPU cannot accommodate all model parameters or large batches of input sequences in onboard memory^30^.

LLMs have been employed previously in biomedical domains. Many LLMs specialized on biomedical domains are trained on publicly available datasets such as abstracts and text from scientific literature as well as deidentified EHR datasets like MIMIC-III^31^. Due to limitations in availabilty of public de-identified clinical datasets, many biomedical LLMs are pretrained on the same clinical corpora and biomedical literature. For example, BioBART, BioMegatron, BioBERT, BioGPT, and Gatortron all contain abstracts and/or full-text from PubMed^32^ in their pretraining dataset, and ClinicalBERT and Gatortron have both seen MIMIC-III. While also trained on scientific abstracts, Forge-bio is notable for being trained on a wider set of abstracts sourced from 6 scientific literature databases. Additionally, the Gatortron-line of models and RadBERT have been fit with in-house clinical data from UF Health Integrated Data Repository (IDR) and radiology reports from Veterans Health Administration (VHA) admissions, respectively.

To make comparisons between biomedical LLM architectures, this study gathered 3 characteristics of their pretraining methodology including: 1) an estimate of the necessary compute for pretraining (6 floating-point operations per parameter per token) by conventions developed by Yin et al.^30^, 2) the ratio between pretraining tokens and model parameter count determining compute optimal training as presented by Hoffman et al.^33^, 3) the context width of the model, i.e., the number of input tokens per sequence allowed by an LLM architecture. These characteristics are estimated to the best of our ability from papers and supplementary material, but it is important to note that LLM literature is often opaque with the exact description of pretraining methods, and pretraining conventions have been rapidly changing within recent years. Table 3 reports these characteristics for models covered in the analysis.

**Table 3.** Pretraining Characteristics Across Biomedical Large Language Models.

|  | model | parameters | tokens | context width | seen MIMIC | reported datasets |
| --- | --- | --- | --- | --- | --- | --- |
| 0 | RadBERT_2m | 109.5 M | 466 M | 512 | No | VHA Radiology Reports |
| 1 | Bio_ClinicalBERT | 108.3 M | 666 M | 512 | Yes | MIMIC-III |
| 2 | BioBART_base | 166.4 M | 6 B | 1024 | No | PubMed Abstracts |
| 3 | BioBART_large | 442.3 M | 6 B | 1024 | No | PubMed Abstracts |
| 4 | BioMegatron_base | 333.6 M | 8.1 B | 512 | No | PubMed Abstracts, PMC Full Text |
| 5 | BioBERT_base | 108.3 M | 28.4 B | 512 | No | PubMed Abstracts, PMC Full Text, BooksCorpus US, English Wikipedia |
| 6 | BioGPT_base | 346.8 M | 14 B | 1024 | No | PubMed Abstracts, PMC Full Text |
| 7 | BioBERT_large | 364.4 M | 28.4 B | 512 | No | PubMed Abstracts, PMC Full Text, Books Corpus US, English Wikipedia |
| 8 | BioGPT_large | 1.571 B | 14 B | 2048 | No | PubMed Abstracts, PMC Full Text |
| 9 | Gatortron_base | 355.3 M | 121.33 B | 512 | Yes | UF Health IRD, MIMIC-III, PubMed Abstracts, PMC Full Text, English Wikipedia |
| 10 | Gatortron_s | 355.3 M | 121.33 B | 512 | Yes | UF Health IRD, MIMIC-III, PubMed Abstracts, PMC Full Text, English Wikipedia |
| 11 | Forge_bio | 1.440 B | 38 B | 2048 | No | SciBert Classified Biology/Medicine (Bio/Med) Abstracts From CORE, OAG, MAG, Aminer, ArXiv, and SCOPUS |
| 12 | Gatortron_medium | 3.913 B | 121.33 B | 512 | Yes | UF Health IRD, MIMIC-III, PubMed Abstracts, PMC Full Text, English Wikipedia |

### Overview of Framework

The proposed framework follows an explanatory sequential design splitting analysis of biomedical LLMs’ latent spaces into quantitative then qualitative assessment as summarized by Figure 1. Quantitative assessment is described as follows:

1. Structured admission-level variables *V* are mapped to unstructured discharge reports *C*.
2. For candidate LLMs *m* ∈ *M*, discharge reports *C* are tokenized then embedded forming corpus manifold *E* per model *m*.
3. For each corpus manifold, admissions are clustered into *k* subsets using K-Means clustering.
4. A Kullback-Liebler Divergence (KLD)-based scoring function is used to measure the amount of information lost per subset when assuming a distribution of variables *V* expected by random subset assignment for admissions occurring during the collection period.
5. A linear model is used to fit scores for each subsetted latent space against the estimated compute needed for pretraining, the token per parameter ratio between their pretraining data and model size, and the context width of the LLMs.
6. Linear models are used to fit scores against performance on classification tasks, derived from structured EHR such as phenotyping and overal mortality prediction, using cluster distances as input to logistic regression classifiers.

**Figure 1.**
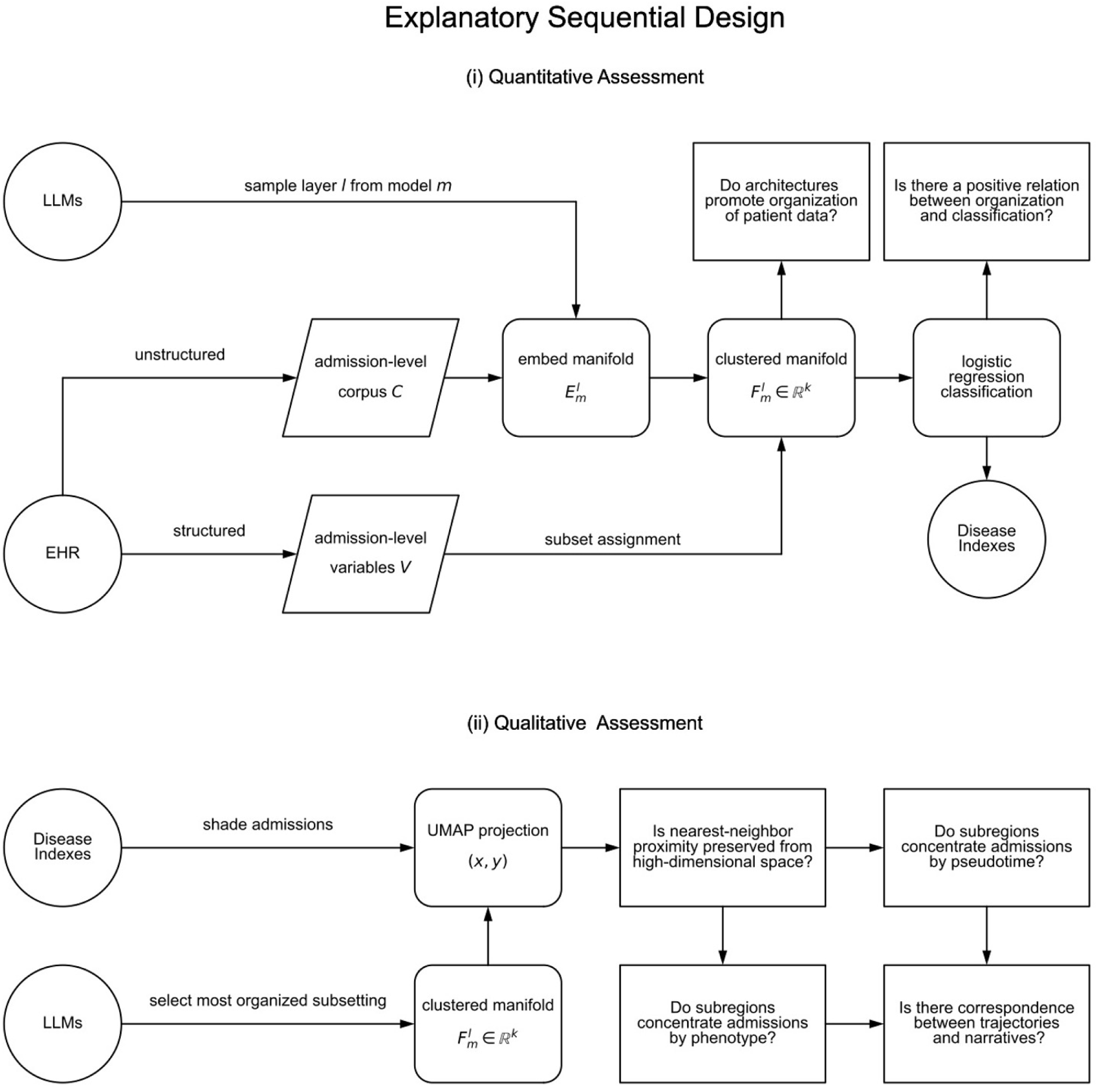
A proposed explanatory framework for evaluating the capability of Large Language Models (LLMs) to represent admission-level clinical text as "maps of disease progression". The framework employs a mixed-methods approach, combining quantitative and qualitative assessments of latent spaces in LLMs. The process starts by mapping structured admission-level variables to unstructured reports, embedding clinical text using various biomedical LLMs, and then partitioning the sampled latent spaces. Admissions are assigned to subsets, corresponding to sub-regions of the latent space. Different classifiers are fit to each intermediate representation to construct indexes of diseases. A qualitative evaluation of these indexes determines the coherence between the trajectories found in low-dimensional projections of the latent spaces and the clinical narratives. This framework aims to provide insights into how diseases and stages of disases are encoded in LLMs, offering a tool to understand disease progression.

After the subsetted latent spaces are scored, the top performing LLM is selected. To investigate applications of biomedical LLM latent spaces for encoding phenotypes and time-dependent health status variables (referred to as "pseudotime" from here on) as well as representing medical trajectories, the top performing LLM is assessed qualitatively for these properties as follows:

1. UMAP^34^ dimensionality reduction is used to plot a 2-dimensional projection of the corpus manifold *E*. Projections are plotted then shaded using color gradients corresponding to a variable of interest *v*.
2. Regions of high organization are observed, shading cluster membership by proximity of clusters’ centroids in the original high-dimensional space and the information surplus over subsets.
3. Temporal ordering is observed, shading projections by time-dependent variables including age at time of admission and time till death at discharge.
4. Phenotypic ordering is observed, shading projections by the prevalence of 6 OSA-related comorbid diseases.
5. Along OSA patients, admissions are plotted then shaded by class probability of comorbid diagnosis and 1-year mortality estimated by logistic regression.
6. Patient admissions are then traced along the corpus manifold and their corresponding discharge reports are reviewed.

### Embedding An Admission-Level Corpus

We refer to embedding a complete set of documents *C* as constructing a corpus manifold *E*. Embeddings for all 331,794 discharge reports in MIMIC-IV were computed using a set of biomedical LLMs *M*. This process involved tokenizing the text and then sampling layer activations for each model *m*.

Without prior knowledge of which specific layer holds the best-compressed representation of the clinical documents, activations were sampled from the last *L* layers per each model *m*. The token-wise MLP activations at the end of a transformer block were collected and averaged into a single vector. If the context width for model *m* was shorter than a full discharge report, the token sequence was chunked into pieces, each of size equal to or less than the maximum context width, without overlap. Considering *C* as an admission-level corpus of the Boston’s Beth Israel Deaconess Medical Center, the transformation *m*: *C* → ℝ*^d^* assigns an admission-level feature vector to each report, where *d* is the number of token-wise features in model *m*. The construction of an admission-level vector per report can be summarized as

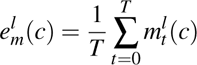

where *l* is the layer being sampled, *t* is the token index along the sequence, and *c* ∈ *C*. The collection of admission-level vectors 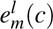 form one corpus manifold 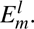 Using an allocation of 290.8 node hours on NERSC’s Perlmutter supercomputer, we sampled corpus manifolds from the last 3 layers of the 13 models covered in Table 3. Data parallel inference was implemented to accelerate construction of manifolds using 32 Nvidia A100 GPUs. These corpus manifolds were used to ascertain the concentration of medically relevant variables along subregions of the LLMs’ latent space as described in the following sections.

### Subsetting Latent Spaces Using K-Means

Analysis of healthcare data often requires the subdivision of patient records into smaller, domain-specific cohorts to focus on particular phenotypes or to establish suitable control groups for case-control studies. The exponential increase in records within EHR databases necessitates scalable techniques for grouping patients based on exposures to environmental factors, occurrences of medical events, and longitudinal outcomes^35^.

To prepare the latent spaces for evaluation, corpus manifolds were initially partitioned into subregions to assign admission-level representations to different groups. Without prior knowledge of the underlying number of potential subgroups present in the clinical dataset, K-Means clustering^36^ was utilized with varied number of expected clusters *k*. Due to constraints in available computational resources, corpus manifolds were subsetted into 256, 512, 1024, and 2048 clusters. For each corpus manifold 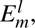 admission-level vectors were regularized using max-norm regularization^37^ then partitioned into *k* sets *S* = {*S*_1_*, S*_2_*, …, S_k_*}. For a given set *S_i_*, the cluster’s centroid along the corpus manifold is described as

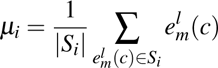

where |*S_i_*| represents the size of the cluster. In addition to assigning each admission to a distinct cluster, the distance relations to each cluster’s centroid were computed to be later used as input for classification models described in the subsequent sections. After centroids are identified with K-Means, vectors on corpus manifold 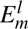 are mapped to cluster-space 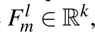 where each dimension in the cluster-space vector corresponds to the regularized distance from the admission-level vector to each cluster centroid found by K-Means.

### Scoring Implicit Ordering in Foundation Models

Often referred to as "foundation" models, the capability of LLMs to transfer features learned during pretraining to problems not explicitly defined in their original training policy extends the concept of transfer learning in ML^38^. The two stages of LLM transfer learning, termed "pretraining" and "finetuning," involve initially training a model on the more general task of token prediction, followed by retraining on a more specific problem with less training data. While transfer learning predates the introduction of LLMs, these models show promise as the first iteration of foundation models due to the apparently tangential yet simple-to-prepare task of pretraining on next-token prediction compared to the domain-specific problems tackled by NLP applications.

Considering auto-regressive architectures exclusively at this stage, the explicit task of language modeling can be expressed using conventions in this paper as follows:

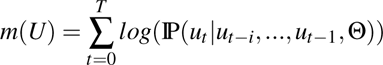

where *m*(*U*) is the model loss over the full corpus of pretraining text and IP(*u_t_*|*u_t_*_−*i*_*, …, u_t_*_−1_, Θ) is the conditional probability of the token *t* given *i* preceding tokens and model parameters Θ^39^. Often referred to as the perplexity of the LLM along the pretraining dataset, it’s difficult to gather these measures for analysis without access to those datasets used to pretrain each model.

Currently, the field of foundation modeling is considered relatively young, and methods for evaluating foundational capabilities vary widely between projects and domains. By "foundational", we imply a common converging cornerstone representation for a specific domain as opposed to the common catch all "foundation" for LLMs trained at scale. While it is common for LLMs to be evaluated using community NLP benchmarks and human-grade standardized tests, several concerns arise due to the opacity in large-scale LLM research to outside institutions^40^. Particularly, it is challenging to assess the level of bias in benchmark construction (such as cherry-picking benchmarks) and data leakage in the pretraining dataset (where tests and answers may be present in the dataset)^41^. Additionally, the cost of curating gold-standard NLP benchmarks is expensive in terms of experts and time needed for annotation^42^. With millions of patients and medical records, curating benchmarks for the field of healthcare is prohibitive.

As opposed to evaluating model capability along a behavioral framework^40^, we propose developing more rigorous scientific frameworks to describe the underlying order found in LLM latent spaces by generating hypotheses for "synthesized designs" solved implicitly during explicit pretraining on next-token prediction, then detecting for the presence of solutions to these hypothetical tasks empirically. We use synthesized designs as a concept that offers information about how practices in foundation modeling lead to organization along hidden representations of contemporary models. If intermediate solutions for these implicit tasks exist within an LLM’s latent space, high degrees of order along these tasks would justify performance seen on downstream finetuning tasks, interpreting high competency on a finetuning problem as indicative of successful encoding of relevant information used in finetuned models along subregions of the pretrained latent space.

In the context of clinical finetuning, we hypothesize biomedical LLMs order admission-level reports into collections capturing distinct clinical phenotypes at different stages of disease. We posit a foundational model for clinical text compresses information seen during pretraining into latent representations encoding “disease states” - subsets of admission-level representations with common phenotype and pseudotime. Transitions between disease states would form trajectories detectable along a corpus of longitudinal medical data. We define a Kullback-Liebler-based^43^ scoring function to measure the amount of information content or "surprise" captured along subregions on an admission-level corpus manifold as follows

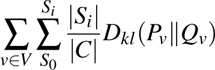

where *V* is a set of medical variables of interest, |*S_i_*| is the size of admission subsets, |*C*| is total number of admissions in the cohort, *P_v_* is the probability of observing the medical variable of interest *v* within a subset, and *Q_v_*is the probability of observing the medical variable of interest *v* along a subsetting which assumes random assignment for admissions occurring during the collection period of the dataset. We consider the demographic determinants of the patient (sex, race, ethnicity), age at time of admission (binned at every 5 years^44^), weeks till death at time of discharge, ICD diagnoses per admission and Current Procedural Terminology (CPT)-coded services as a candidate set of medical variables *V* necessary to encode a disease state. For ICD and CPT variables, codes were clustered to reduce each set of codes to 100 degrees of freedom for numerical stability. Used as a scoring function for biomedical LLM latent spaces, the measure indicates the amount of information lost per subset when assuming subsetting contains no discernable discrimination between the local statistics of subsetted admissions and the global statistics of admissions in the dataset.

As a consequence, we posit the proposed scoring function will inform progress towards a "clinically-aware" design. Higher scores indicate greater deviation from random assignment of subsets, suggesting a more structured arrangement of admission-level representations is present within latent spaces. Higher scores may also suggest that models effectively learn to differentiate between distinct clinical observations localized by time, indicating intermediate solutions towards modeling longitudinal outcomes and serving as a strong foundation for downstream clinical tasks in predictive medicine. Conversely, lower scores may indicate challenges in capturing meaningful structure within subregions of the latent space, highlighting areas of high uncertainty where further model refinement should be done through supplementation with additional clinical observations.

### Admission-Level Classification Using Clusters Distance

To evaluate the hypothesis posited in the previous section, the performance on admission-level classification tasks was assessed for each corpus manifold. If the hypothesis holds true, a positive relationship would be expected between the score of the corpus manifold on the previously described implicit task and the performance of classifiers on explicit phenotypic and pseudotime tasks.

Cluster-space vectors 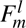 for each corpus manifold were utilized as input to classify admissions from patients with history of OSA. Each admission was labeled by the diagnosis of OSA and its six associated comorbid diseases. Furthermore, these admissions were labeled by outcomes for overall mortality used to predict death 1-year, 90-days, and 30-days after discharge. Logistic regression with Elasticnet regularization^45^ was employed, using a combination of L1 and L2 penalties to prevent overfitting and improve model interpretability with respect to which regions of the latent space contribute to classification. Without prior knowledge of the needed complexity for each linear model, the regularization coefficient was set to a value of 1.0 and the L2 penalty term was set to 0.5 consistently across all classifiers per corpus manifold.

Logistic regression classifiers were fit on the 10 binary classification tasks per corpus manifold. K-fold validation on each classifier was conducted, yielding the mean area under the ROC curve (AUC-ROC) over 16 folds. This approach ensured robust evaluation of classifier performance across different subsets of the dataset as per-classifier heuristic optimization is absent at this time. While more optimal classifiers can be constructed with hyperparameter search for each tasks, measuring the difficulty of finding performant classifiers from intermediate solutions on the corpus manifold is the goal of this technique.

### Dimensionality Reduction Using UMAP

Given the high-dimensionality of the admission-level embeddings, we explore employing the dimensionality reduction algorithm UMAP to generate human-readable 2-dimensional plots of corpus manifolds for MIMIC-IV. The UMAP algorithm is known for preserving local structure between points while also maintaining more global structure compared to other dimensionality reduction methods like PCA^46^ and t-SNE^47^. This property renders UMAP a valuable technique for interpreting encodings or embeddings of high-dimensional data and has been successfully applied in previous work to visualize biological datasets such as transcriptomes of single-cell data^48^.

In the context of visualizing a corpus manifold 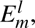 each admission-level vector is mapped to a coordinate (*x, y*) on a 2-dimensional plane. Each point in the resulting projection corresponds to a single admission within the healthcare system. Through a grid search over UMAP parameters (such as number of nearest neighbors per high-dimensional point and minimum distance between points in the projection), an adequate projection of the corpus manifolds was attained, neither tightly-packing points nor dispersing points uniformly. However, it is important to note that there currently lacks a standardized method for improving the quality of the projection, often relying heavily on trial and error. Additionally, distances along the projection do not necessarily imply meaningful relations between points, but rather denote neighborhood proximity, as is also observed with t-SNE. The cosine distance between admission-level cluster-space vectors 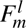 were used to fit the UMAP projection.

### Chart Review For Cases of OSA Patients With Concurrent HF

As a case study in using biomedical LLMs to characterize severe disease phenotypes, a chart review of discharge reports for patients with history of OSA was performed to assess if comorbid diagnosis probability along the corpus manifold can be used to index severe illness, and if mentions of severe OSA are enriched among regions of the corpus manifold with high probability of HF diagnosed admissions.

Matched stratified sampling was used in this study. OSA patients with concurrent HF were matched with OSA patients with no history of HF along sex, race, and age at time of their first recorded diagnosis of OSA. After cases and their controls were identified, admissions with similar age at time of admission (within a 5 year window) were gathered and 1 admission per case-control match were randomly sampled. This was done as patients potentially have multiple admissions at varied times over the collection period of the dataset. Its important to note that multiple rounds of chart review were conducted to update exclusion criteria for admissions including: 1) removing non-relevant specialties like ‘OBSTETRICS/GYNECOLOGY’ and ‘ORTHOPAEDICS’, and 2) removing ‘ELECTIVE’ admission types.

74 admissions were considered in the final chart review where reviewers were tasked with annotating evidence for severe OSA (prolonged hypoxia or respiratory obstruction) as a mechanistic driver for HF including mentions of: 1) respiratory distress such as hypoxia, dyspnea, shortness of breath, or related symptoms, 2) history of diagnosed OSA in the patient’s medical history preceding current admission, 3) prescribed OSA treatment, such as CPAP devices or specific pharmacological interventions, and 4) adherence/compliance to OSA treatment. After annotation was complete, the HF class probabilities were merged for analysis to measure the odds-ratio of severe OSA or poorly managed OSA over strata of HF probability.

## Results

Table 1 reports the cohort characteristics covered by the MIMIC-IV discharge report corpus. The cohort consists of 145,915 patients, with a close to even distribution of sex at 48.7% male and 51.3% female. The majority of patients (68.6%) identify as White, followed by Black (11.98%); 4.8% of patient identifying as Hispanic. Notably, 18.7% of patients in the cohort are marked as deceased with the majority of deaths occurring between the age 60 and 90. The majority of admissions were categorized as emergency (87.8%) with the remaining considered as elective (12.2%). The average number of patient-days summarized by a single discharge report is 4.92 patient-days.

A total of 14,327 patients were diagnosed with OSA with increased rates of males along sex, increased rates of whites and blacks along race, and no discernable change in Hispanic ethnicity when compared to the overall population. While overall mortality drops marginally when compared to non-OSA patients, OSA patients have much higher rates of the 6 comorbid diseases than non-OSA patients. For OSA patients, age at time of admission tends towards an older population and shorter lifespans for those patients with reported mortality outcomes. Subsequent sections are divided into results for quantitative and qualitative assessments respectively followed by results from the chart review performed on OSA patients with concurrent HF.

### Quantitative Assessment

#### Subsetting Clinical Admissions By Phenotype and Pseudotime

First, a linear regression analysis was conducted to investigate the potential positive relationship between the three architecture characteristics gathered for each biomedical LLM and the surprise in observing implicit ordering by disease state, as defined in the previous sections. Considering the best layer and subsetting of the corpus manifold per model, the regression model indicates strong evidence that increased compute, compute-optimal training, and a widening context correlate with order among subsetted latent spaces for our proposed implicit task. In other words, the regression model suggests that 60.3% of the variance on the proposed task can be explained by decisions made at the architecture level.

Upper and lower bounds (95% CI) of compute-optimal ratios and context width help explain the underperformance of certain models. Specifically, models with short context widths (512 tokens) underperform even when approaching compute-optimal training, as seen in the publicly-available Gatortron-line models. Interestingly, plotting the trendline for compute-optimal training at the maximum context-width in this sampling of biomedical LLMs retrodicts the field’s adherence to compute optimal conventions. Both the large variant of BioGPT and Forge-bio reported results for their pretraining (October 2022 and November 2023, respectively) after the preprint publication of Hoffman et al.’s paper (March 2022). Gatortron reported results for pretraining in February 2022, one month before Hoffman et al.’s paper was posted on ArXiv. Results for this regression analysis are plotted in Figure 2a.

**Figure 2.**
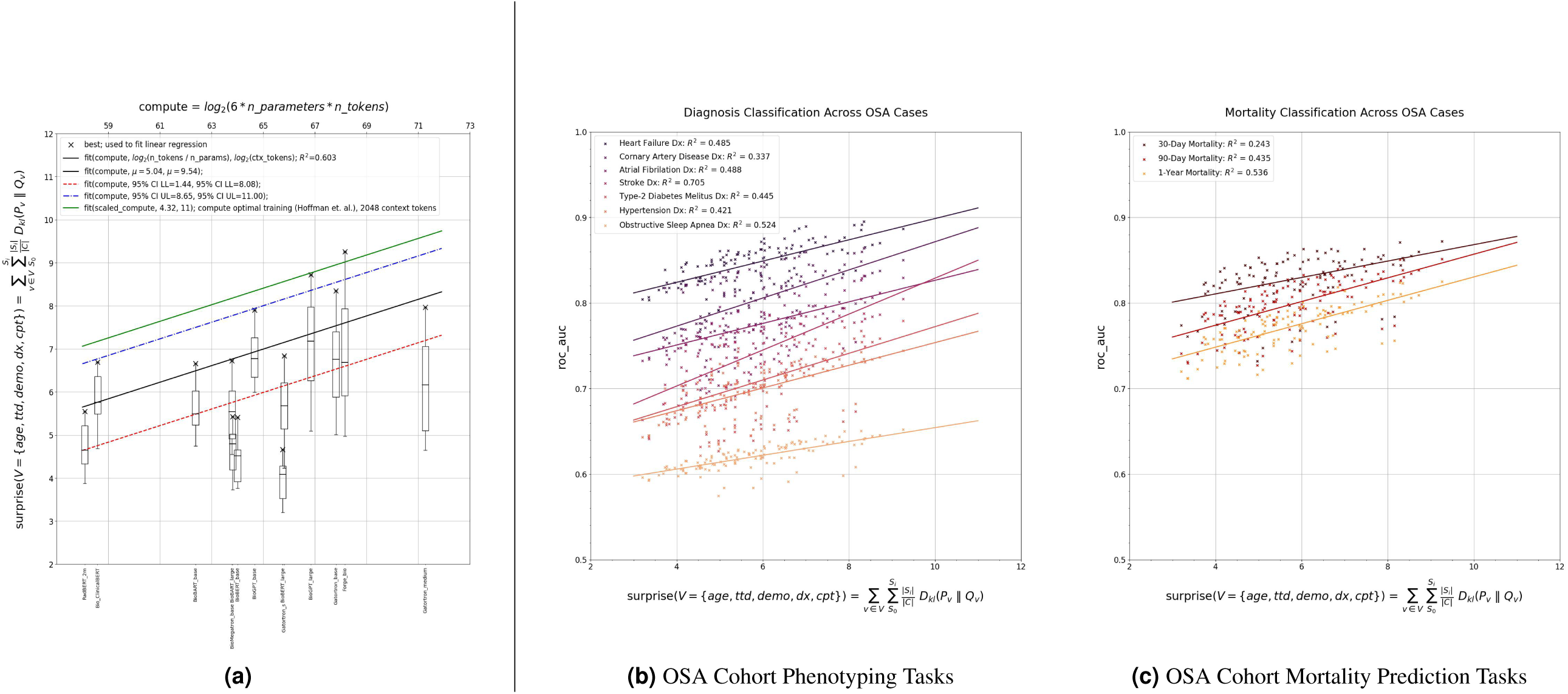
Quantitative results across biomedical LLMs covered in the study. **(a)** Plots subsetted latent space surprise over the estimated compute needed to pretrain each model with compute optimal conventions reported by Hoffman^33^ and Yin^30^. Box plots show the distribution of scores per model over different layers and subsettings. Compute, ratio between training tokens and parameter count, and context length of pretrained models explain 60.3% of variance when regressing against scores. **(b)** Plots area under the ROC curve (ROC-AUC) performance on admission-level phenotyping tasks over subsetted latent space scores. Moderate to strong relationships are found between performance on tasks and scores. **(c)** Plots ROC-AUC performance on admission-level overall mortality prediction over subsetted latent space scores. Moderate to strong relationships are found between 90-day and 1-year mortality prediction and scores.

Table 4 reports more granular measures over the top 10 subsetted corpus manifolds, with Forge-bio containing the highest amount of surprise overall and across age at time of admission, weeks till death after discharge, ICD diagnosis, and CPT services individually. Along demographic determinants, the fully-trained baseline Gatortron model marginally outperformed Forge-bio. This may be a result of observing clinical data, but there are no other models of comparable size trained on clinical data in this work to confirm that pretraining on clinical data allows for better discrimination of demographic determinants at this time.

**Table 4.**
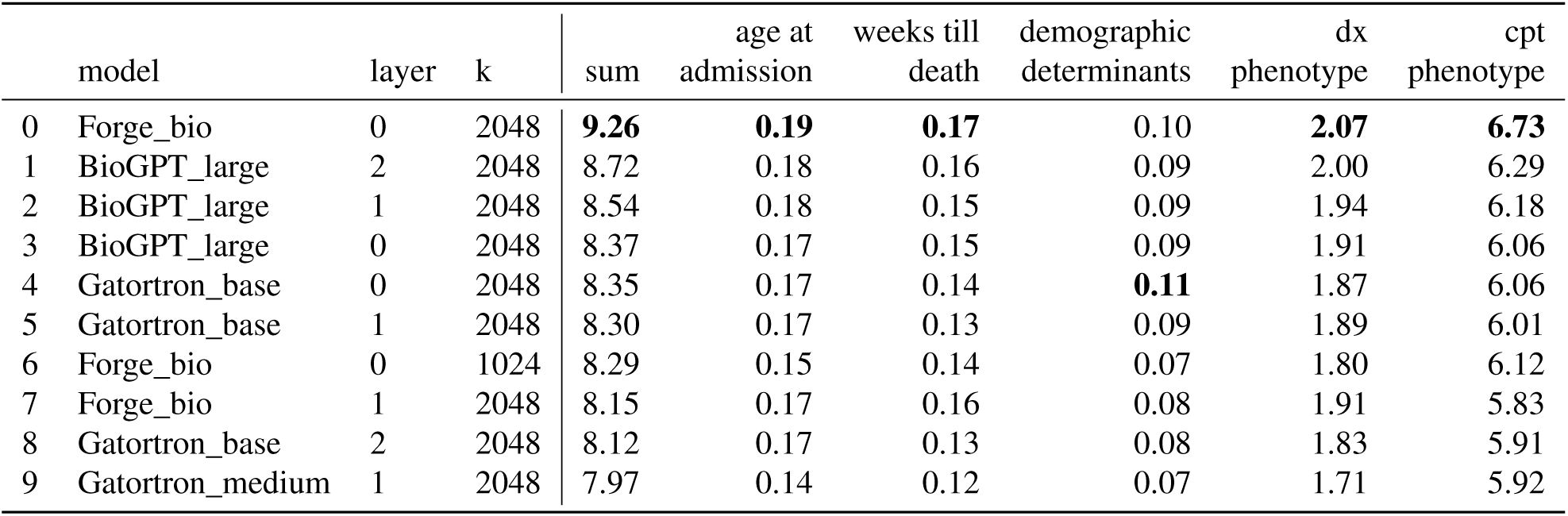
Top Subsetted Latent Spaces by Proposed KLD-based Scoring Function.

Second, two linear regression analyses were conducted on the potential positive relationships between the surprise in observing implicit ordering by our proposed definition of disease state and the phenotypic/pseudotime tasks in the OSA case study. Over all subsetted corpus manifolds sampled from the 13 biomedical LLMs, the regression models indicate moderate to strong evidence that implicit ordering by disease state helps provide adequate intermediate solutions for classifiers on the comorbid disease classification and overall mortality prediction tasks. Performance on overall mortality classification by AUC-ROC decreases as the time window widens from 30-day to 1-year mortality, with only weak evidence for correlation between surprise and mortality prediction 30 days out.

Along phenotypic tasks, there is strong evidence of correlation between surprise and improvements in diagnosing admissions for STRK and OSA. Even so, OSA classification only showed marginal improvements as a function of surprise when compared to STRK. Moderate evidence was found for HF, CAD, AFIB, T2DB, and HTN, with HF classifiers performing highest along AUC-ROC. Given that this sampling of classifiers is not heuristically optimized, the AUC-ROC scores reported along these results are not conclusive on the limit of performance for any one classification problem, but we expect increasing the computational budget to accommodate for hyperparameter search will help give better assessment on the limits of performance per task while still indicating a positive correlation between surprise and discriminatory power. Results for these two regression analyses are plotted in Figures 2b and 2c.

### Qualitative Assessment

For the best subsetted corpus manifold (last layer of Forge-bio subsetted into 2048 clusters), Figure 3 plots the UMAP projection of the corpus manifold. First, the subset assignment for each admission is shaded by cluster, with the color gradient indicating proximity between the cluster centroids. While no clear separation is found to indicate independent groups of admissions, the UMAP projection does preserve neighborhood proximity found in the higher-dimensional space with bands of admissions expressing similar concentration of colors on the gradient. The overall structure of the projected corpus manifold resembles a curved Y-shaped band centered at *y* = 0.55. A smaller independent patch can be seen towards the bottom left of plot, centered at (0.25, 0.20), and a small thin Y-shape band at *y* = 0.7. Second, plots are shaded by the per-cluster surprise, with the most order following the core of the large Y-shaped band. One point of contention in interpretation of UMAP plots is the difficulty of discriminating what structures in the projections are artifacts produced as side-effect of the algorithm. Even so, we observe enough preservation of the high-dimensional space to proceed with interpreting results for ordering admissions by phenotype and pseudotime.

**Figure 3.**
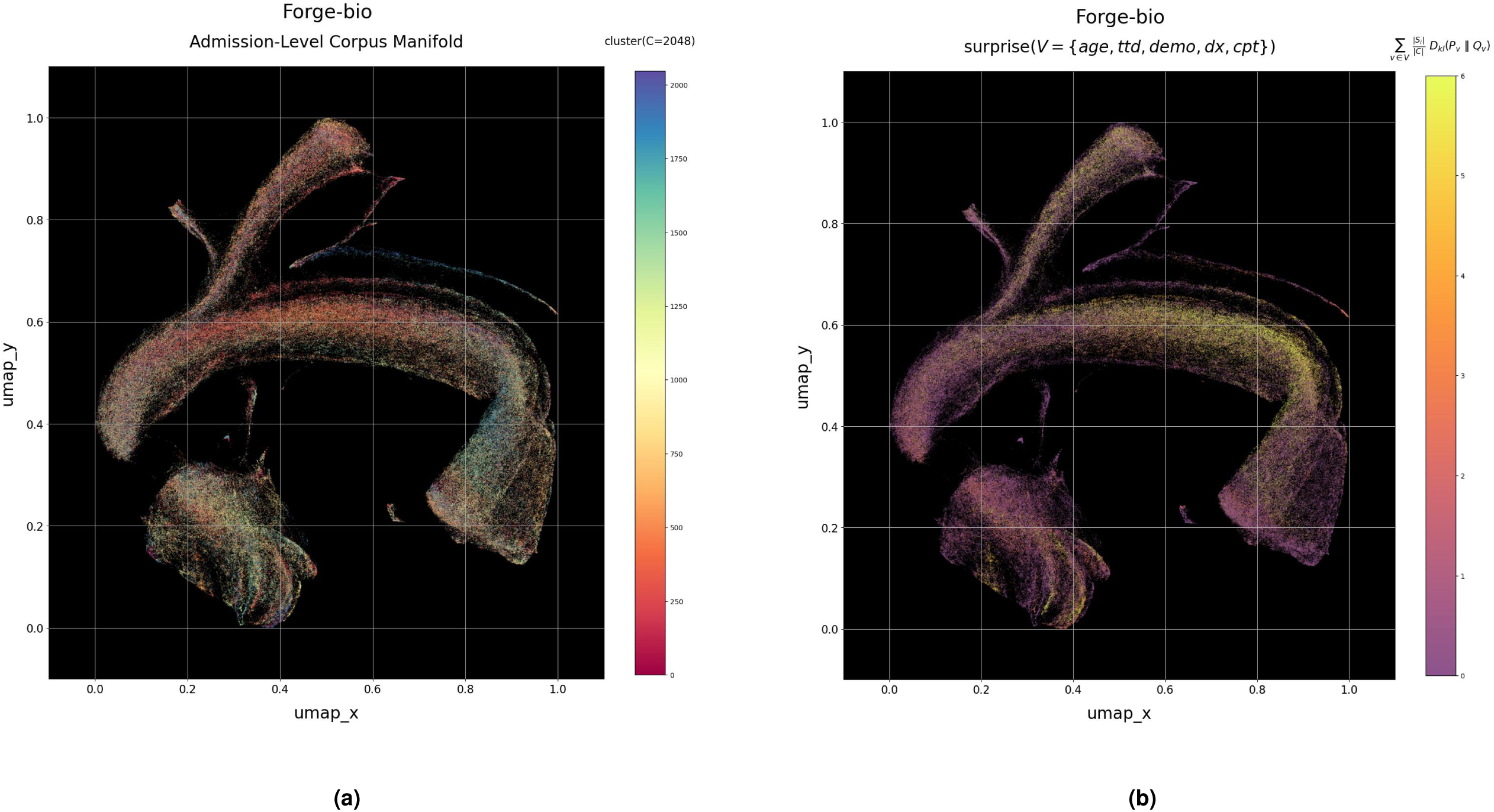
Qualitative evaluation of best subsetted latent space (Forge-bio, k=2048, penultimate layer). **(a)** UMAP visualization of admission-level corpus manifold. Color gradient corresponds to proximity of K-Means clusters. **(b)** Per cluster KLD scores over corpus manifold. Intensity corresponds to higher amounts of order.

#### Ordering By Pseudotime Variables

Figure 4 is divided into two sections, with the first shading the average age at time of admission per subset. Notably, the projected Forge-bio corpus manifold show regions with concentrations of admissions with patients 35 years old and younger centered at (0.175, 0.175). The small thin Y-shaped band referenced in the previous section also concentrates admissions with relatively younger patients (< 40 years). Concentrations of admissions with patients 65 years and older can be seen along the core of the large Y-shaped band. The second section of the figure shades time till death after discharge by prevalence of 1-year, 90-day, and 30-day mortality per admission subsets. As shading moves towards 30-day mortality, we can see recession of prevalence along the large Y-shape band with the strongest concentration remaining around the region at [(0.1, 0.2), (0.4, 0.5)]. Interestingly, the thin section within that region, at the top of the independent patch at the bottom left of the plot, has overrepresentation of deaths occurring during admissions to the ICU. In conjunction, these two views demonstrate the temporal ordering of admissions in MIMIC-IV by age and life expectancy. However, its important to note that MIMIC-IV is temporally sparse along longitudinal data and is predominently acute emergency admission to the ICU. A larger longitudinal dataset is needed to qualitatively assess if these observations persist along wider windows of time.

**Figure 4.**
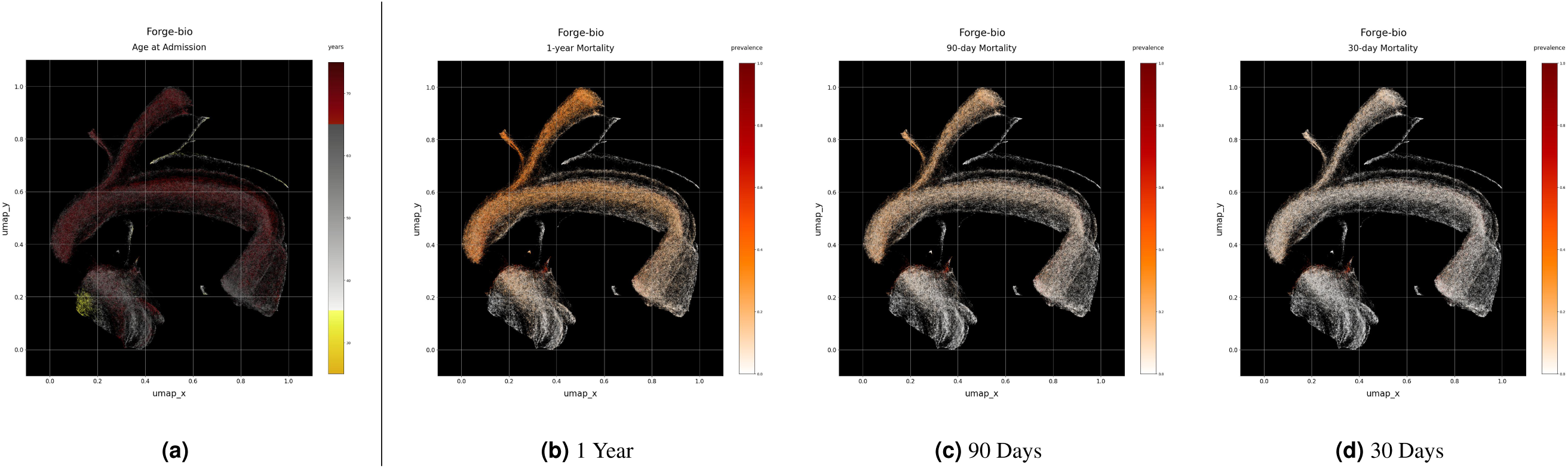
Prevalence of time-dependent variables over clusters on corpus manifold. **(a)** Plots mean age of patients at time of admission. Yellow regions correspond to clusters with patients 35 years-old and younger. Red regions correspond to clusters with patients 65 years-old and older. **(b,c,d)** Plot 1-year, 90-day, and 30-day overall mortality respectively. High prevalence clusters for overall mortality concentrate around center-left region at [(0.1, 0.2), (0.4, 0.5)].

#### Ordering By Phenotypic Variables

Figure 5 visualizes the prevalence of the 6 comorbid diseases of interest for the OSA case study. Visualizations for prevalence of HTN and T2DB show moderate to high prevalence over the large Y-shaped band, the small thin Y-shape band and the independent patch at the bottom left of the plot. Prevalence recedes over subregions that were previously referenced as having higher concentrations of admissions for patients age 40 years and younger. The prevalence of AFIB and CAD concentrate mainly along the large Y-shaped band, with increased concentration on the top of horizontal portion of the band at *y* = 0.6125. Interestingly, this Forge-bio corpus manifold does show clear separation between regions with high concentrations of HF and STRK. STRK admissions are found in abundance following thin bands at *y* = 0.675 and *y* = *x* + 0.225. For HF, there is heavy overlap with regions concentrating AFIB and OSA. Clear separation of the STRK phenotype may help in interpreting the sharp increase in classification performance seen during quantitative assessment.

**Figure 5.**
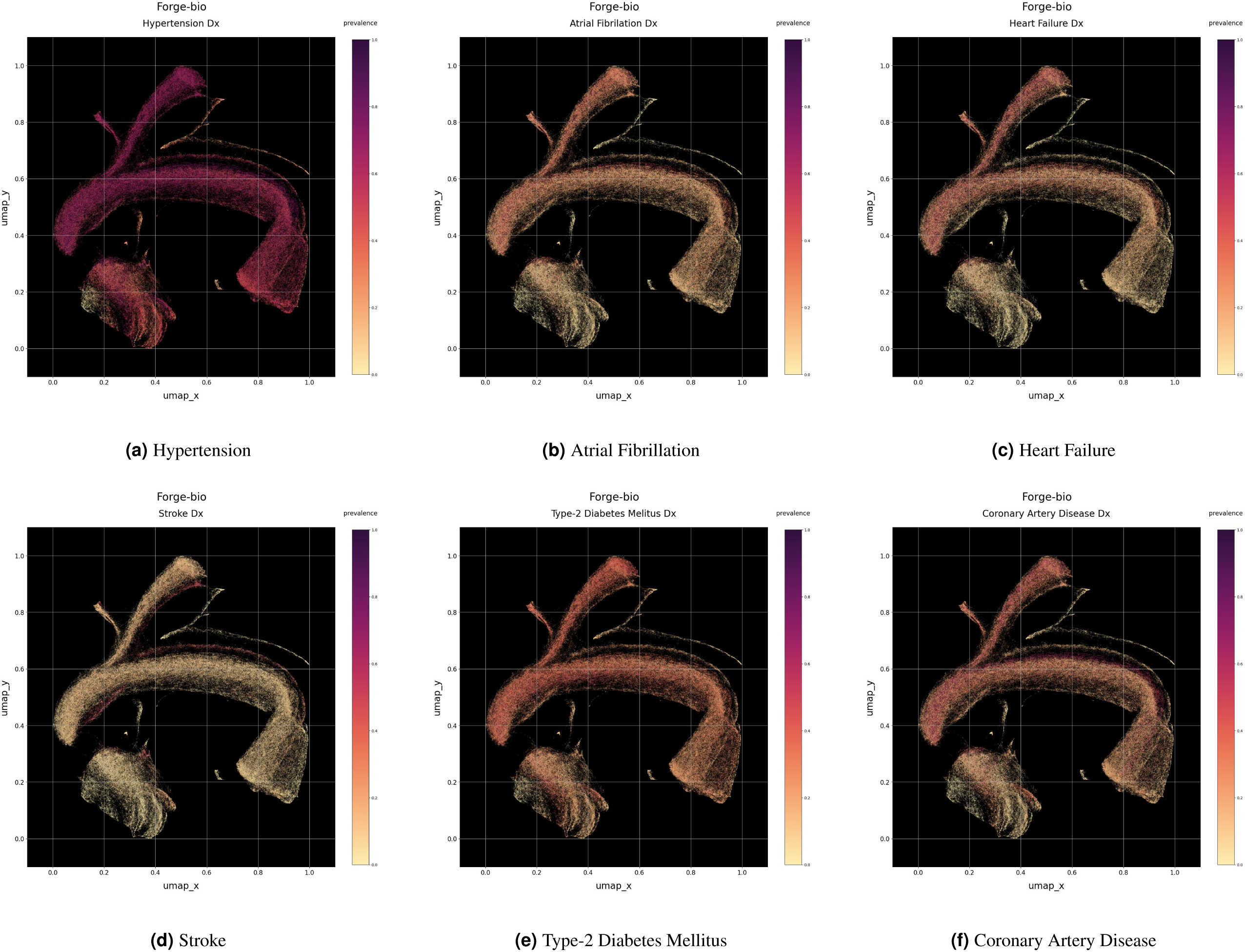
Prevalence of 6 OSA-related comorbid diseases over clusters on corpus manifold. **(a, e)** HTN and T2DB are prevalent across manifold with reduced prevalence observed along clusters with younger average age at time of admission. **(b, f)** AFIB and CAD concentrate on clusters along *y* = 0.6125. **(c, d)** Seperation between HF and STRK is observed. STRK concentrates on clusters following the bands at *y* = 0.675 and *y* = *x* + 0.225. In contrast, HF overlaps heavily on regions with concentrations of AFIB and CAD.

#### Trajectories Along Corpus Manifolds

Admissions for OSA patients with concurrent HF were traced through the corpus manifold to interpret how regions of the projected manifold correspond to narratives found in the discharge reports. Patients whose trajectories terminate as deaths during an admission were selected. Figure 6 visualizes admissions along the Forge-bio corpus manifold for patients with diagnosed OSA. Plots on the left column shade the class probabilities of HF diagnosis from the logistic regression model trained during quantitative assessment. Plots on the right column shade the class probability for 1-year Mortality. Each row in the figure corresponds to a unique OSA patient where admissions with either OSA or HF diagnosis are represented by a cross. Starting at the admission with the earliest recorded diagnosis of OSA, a numbered sequence is traced between admissions to determine if episodes of HF can be inferred by transitioning from regions with low class probability for HF to high probability regions, and if migration towards areas of high-class probability for overall mortality occur as admissions get closer to time of death.

**Figure 6.**
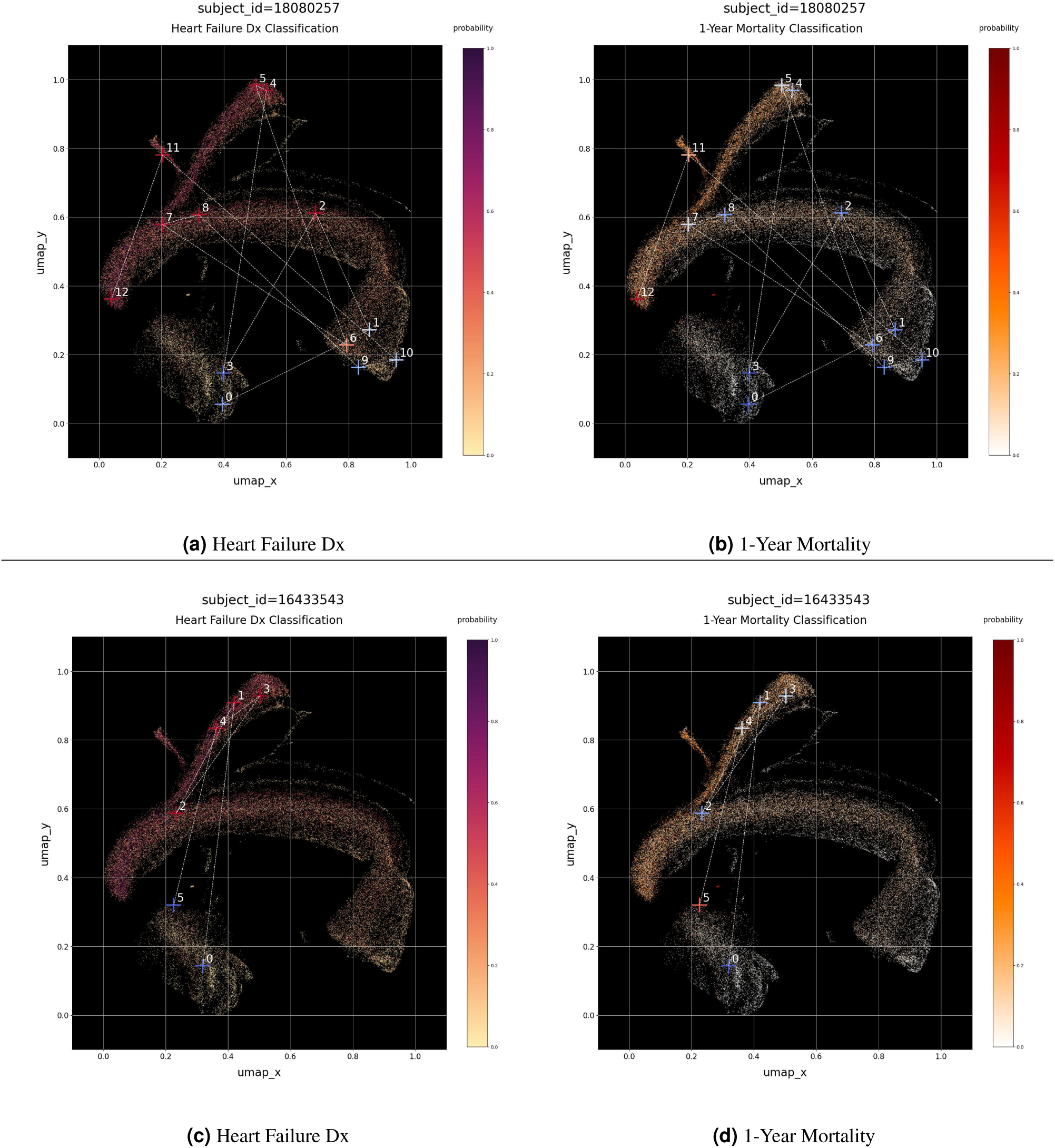
Admissions for two expired OSA patients with concurrent HF traced over corpus manifold. Plots class probabilities for HF diagnosis and 1-year mortality along cases of OSA (51, 565 *admissions*) using logistic regression fit on K-Means cluster distances. Crosses represent unique admissions and thier color gradient correspond the admissions’ class probability. **(a, b)** Plots follow admissions with diagnosed HF or OSA for *sub ject*_*id* = 18080257. Beginning at the earliest admission with OSA diagnosis (*hadm* = 0), 3 episodes of HF can be inferred at *hadm* = 2, *hadm* = (4, 8), and *hadm* = (11, 12). 1-year mortality probability increases with each episode, ultimately terminating with the patient’s in-hospital death at *t* = 12. Chief cause of death, congestive HF. **(c, d)** For *sub ject*_*id* = 16433543, only 1 episode of HF can be inferred at *hadm* = (1, 4). 1-year mortality probability has a sudden increase at *hadm* = 4 with the patient dying in the following admission (*hadm* = 5) from non-traumatic intracerebral hemorrhage.

Consider the first patient’s traced admission history (subject_id=18080257). We see 3 potential episodes of HF along their path through the corpus manifold summarized in discharge reports as follows:

- **First Episode**: Initially, 6.17 years before death (*hadm* = 0), the patient, suffering from comorbid AFIB/CAD and on anticoagulant medication, underwent an uneventful orthopedic surgery with no noted complications. The 4.86 years before death (*hadm* = 1), the patient underwent another orthopedic procedure, tolerated it well, and resumed previously prescribed medications. About two months later (*hadm* = 2), the patient began experiencing intermittent chest pain, which occasionally woke him from sleep and was associated with dyspnea.
- **Second Episode**: Approximately 4.19 years before death (*hadm* = 3), the patient experienced significant right shoulder pain following a proximal humerus fracture repair. Two years later (*hadm* = 4), the patient suffered a heart attack necessitating stenting of a coronary artery, complicated by a groin hematoma. A little over four months after that (*hadm* = 5), the patient presented with a fall, hypoxia, and angina, treated with sublingual nitroglycerin and antibiotics for suspected pneumonia. Another 4 months pass (*hadm* = 6), the patient experienced another cardiac event with mild troponin elevation. About 1.53 years before death (*hadm* = 7), the patient was hospitalized for severe bleeding and hypotension, complicated by a non-ST elevation myocardial infarction (NSTEMI) requiring intensive care. Only 23 days later (*hadm* = 8), the patient presented with rectal bleeding and unstable angina, with recurrent chest pain indicative of non-ST elevation myocardial infarction amidst ongoing complex medical management.
- **Third Episode**: Just under 1 year before death (*hadm* = 9), the patient was admitted following a fall, complicated by a small subarachnoid hemorrhage and reversal of INR. Thirty-three days before death (*hadm* = 10), the patient required surgery for a periprosthetic joint infection of the right shoulder, managing postoperative pain and progressing well. Four days later (*hadm* = 11), the patient returned with bleeding from the surgical site, complicating severe cardiac issues including a non-ST elevation myocardial infarction (NSTEMI) and acute HF with pulmonary hypertension and reduced ejection fraction. His condition deteriorated further with a subsequent stroke and ongoing fluid management challenges. Six days before death (*hadm* = 12), the patient experienced a life-threatening rectus sheath hematoma requiring embolization, followed by cardiac arrest with successful resuscitation but subsequent multi-system organ failure and decision for palliative care. Chief cause of death was recorded as congestive HF.

Next, consider the second patient’s traced admission history (subject_id=16433543). Only 1 potential episode of HF is seen along their path through the corpus manifold, ultimately terminating at a different subregion than the previously described patient.

- **First Episode**: Starting 5.82 years before death (*hadm* = 0), the patient underwent a radical nephrectomy without complications. 7 months before death (*hadm* = 1), the patient presented with acute HF with reduced ejection fraction (HFrEF), managed with diuresis and cardiac catheterization showing non-contributory coronary lesions. The patient was noted to have an underlying sleep disorder. Two months later (*hadm* = 2), worsening dyspnea, leading to a diagnosis of HF attributed to hypertension and potential sleep disorder or stimulant medications. Ninety days before death (*hadm* = 3), recurrent falls and declining functional status prompted hospitalization. OSA was referenced as an acute active issue. Sixty-four days before death (*hadm* = 4), the patient was readmitted for acute-on-chronic HF exacerbation. The final hospitalization (*hadm* = 5), ending in death from non-traumatic intracerebral hemorrhage, involved a decision to transition to comfort measures only (CMO) due to irreversible neurologic decline and poor prognosis.

### Characterizing Drivers of Severe Illness in Obstructive Sleep Apnea

Manual reading of discharge reports was conducted by reviewers (n=2) to observe the concentration of mentions for severe symptoms of OSA over admissions indexed by the probability of HF diagnosis. The objective of the chart review was to investigate whether these indexes enrich evidence for the relationship between indicators of severe or poorly managed OSA and concurrent HF. Multiple rounds of chart review were conducted to ensure consensus between reviewers, using Cohen’s Kappa statistic^49^ to assess inter-rater agreement along 4 categories of annotation. Over the 74 admissions covered in the final chart review, the reviewers had substantial agreement (0.734 - 0.795 Cohen’s Kappa) when annotating for the presence of dyspnea, previous history of OSA at time of admission, and evidence of treatment adherence. Near-perfect agreement (0.858 Cohen’s Kappa) was attained when annotating presence of OSA treatment.

Considering a threshold of 0.52 along HF probability as the decision boundary for admission-level diagnosis, classification of HF diagnosis over the 74 admissions results in a PPV of 0.926 and recall of 0.658. For those admissions classified as HF, a 427% increase in the odds of admissions referencing dyspnea (95% CI, [1.71, 17.33]) and a 245% increase in the odds of admissions referencing ongoing treatments for OSA (95% CI, [1.16, 10.75]) were found. Less conclusive results were found for enrichment of mentions of pre-existing OSA and non-adherence of OSA treatment showing a 67% (95% CI, [0.36, 10.75]) and 42% (95% CI, [0.26, 7.34]) increase, respectively.

The findings align with the medical hypothesis that OSA exacerbates cardiovascular comorbidities, specifically HF. As surrogates for high doses of respiratory obstruction caused by OSA, the 4 indicators suggest an increase in prolonged exposure of severe OSA among patients with concurrent HF. Dyspnea, a direct symptom of respiratory obstruction, significantly correlates with HF diagnosis. The modest increase in pre-existing OSA suggests that persistence of OSA symptoms may contribute to HF progression, but a wider longitudinal review is needed. Even with increased active management of OSA through CPAP or supplemental oxygen treatment, non-adherence to OSA treatment underscores the critical role of consistent management in mitigating OSA’s cardiovascular impact. However, a larger patient pool is necessary to determine the reliability of this indicator for high doses of persisting respiratory obstruction given only 9 patients referenced non-adherence.

## Discussion

A wealth of clinical predictive models apply ML methods on structured data from EHR to help physicians and decision makers integrate large amounts of medical information. Even with the increased push towards automated triaging^50^ over the last few years, most operationalized models do not fully utilize unstructured clinical text which often hold rich details of patient health status over time. LLMs are trained at scale on broad data with a lower need of data cleaning and curation. Often termed foundation models, we posit LLMs act as central, unifying storage, an impressive hash system that allows to efficiently organize massive amounts of data into highly compressed representations. Once pretrained, they can be finetuned for a variety of transfer-learning tasks using significantly smaller amounts of data. We believe foundation models have the potential to capture a full range of associations within clinical corpuses, distill that information into multifaceted dense representations, and integrate diverse modalities of data in medicine. Basic research into the representative capacities of pretrained LLMs for key health indicators is necessary before wide-deployment of models into clinical practice, and insights into how information is stored in LLMs may lead to more informed strategies for solving domain-specific clinical tasks.

### Limitations of Analysis

In conjunction, we believe the three linear regression analyses offer researchers in computational medicine a working theory on how the *complexity of LLMs*, influenced by engineers’ architectural decisions, determines the pretrained models’ capacity for solving downstream clinical tasks. Such a theory would be invaluable for deciding which architecture to use, when to reproduce pretraining on larger datasets, justifying the scale of pretraining over clinical text of healthcare systems, and identifying which classes of clinical tasks are likely to perform well by finetuning.

Several limitations persist with this analysis. First, the architectural characteristics are our best estimations from literature and supplementary materials. Second, there is a lack of understanding regarding the added explanatory power from considering the composition of the pretraining datasets, although Forge-Bio’s performance suggests that a more diverse dataset produces better results. Additionally, the Gatortron line of models give some weak evidence of improved discrimination of demographic determinants when ingesting larger amounts of clinical text. Third, the sparsity of MIMIC-IV in terms of longitudinal data makes it difficult to ascertain the consistency of trajectories in latent spaces. More work is needed to determine at what scale LLMs begin to encode trajectories between subregions of the latent space. Lastly, this analysis does not consider any LLMs pretrained on general-domain corpora, though the interest in applying general-domain LLMs in medicine warrants further investigation.

There exists an opportunity for new information retrieval techniques like Retrieval-Augmented Generation (RAG)^51^ to expedite the process of annotation for chart reviews. While the reported chart review finds some evidence for using textual mentions as surrogate indicators for high dosages of severe OSA, inconclusive results over treatment non-adherence highlights the need for methods to quickly automate the annotation of large patient pools to increase the sensitivity of exposures mentioned in clinical text^52^. Even so, this demonstration shows an early attempt for how clinical experts may use the proposed framework for testing medical hypotheses then develop domain-specific finetuning with insights derived from those investigations.

### Future Work, Challenges and Potential

#### Theoretical vs Empirical Latent Space Analysis

A continued debate persists regarding the relationship between metrics in embedding spaces, concept localization, and semantic relations. Notably, language embedding serves as a demonstration of how vector representations can infer analogies (e.g., king - man = queen - woman), yet the extent of these properties remains an open problem in deep-learning-based NLP^53^. Several attempts have been made to construct a semantic calculus out of token-level representations^54,55^, leveraging learned invariants in an embedding space (e.g., (store (not (gross food))) = restaurant). However, it remains unclear whether these invariants are globally consistent across an embedded manifold and how model pretraining influences their instantiations. The introduction of LLMs further complicates the issue, as they demonstrate that many artifacts found in smaller models disappear simply by increasing the number of trainable parameters and the size of the training corpus.

An alternative explanation of how these systems represent language posits that LLMs function as high-capacity locality-sensitive hashes^56^. In essence, LLM parameters serve as a compression of the pretraining corpus. When predicting the next word in a sentence, these models may execute a complex hierarchical lookup in an address space learned through stochastic gradient descent. While the mechanics behind how these address spaces self-organize remain opaque, an empirical framework like the analysis proposed in this work may serve as a sufficient first step in testing LLMs for clinically relevant representations along this line of reasoning. We believe progress in contemporary artificial intelligence research through its persisting and homogenizing coventions^57^ warrant the development of new empirical frameworks towards detecting *synthesized designs*.

The proposed approach is empirical in nature due to its reliance on real-world data and observable outcomes. By scoring the underlying organization of LLM latent spaces with respect to observational data, researchers can empirically evaluate the model’s ability to encode clinical phenotypic markers and mortality-related pseudotime. This stands in contrast to theoretical approaches, which may rely more heavily on abstract formulations or theoretical assumptions about the structure of clinical language in implementing bespoke retrieval methods. Through empirical analysis, researchers can gain practical insights into the capabilities and limitations of LLMs in capturing clinically relevant information within their latent spaces and identify domain-specific areas that need additional refinement as well as formulating new medical hypotheses.

#### Scaling Framework to High-Resolution Population-Wide Longitudinal Datasets

Our ultimate goal is to assess the utility of pretrained latent representations for modeling longitudinal trajectories of patients. We demonstrate how measuring excess self-information over subregions of the latent space can effectively detect systematic increase or decrease in organization concerning phenotypic variables such as diagnosis and healthcare services, as well as time-dependent variables like patients’ age and life expectancy. However, the analysis over MIMIC-IV provides weak evidence for trajectories due to the sparsity of admissions per patient. We anticipate observing stronger temporal ordering in a larger population study with higher rates of healthcare utilization.

A positive finding would reveal corpus manifolds expressing *attractors*, where patients’ embedded clinical histories trace paths through different phenotypic clusters, ultimately terminating at end-of-life phenotypic clusters (e.g., Major depressive disorder -> alcohol abuse -> death by liver failure). If successful, this analysis will serve as a valuable stepping stone toward understanding how clinical LLMs organize information, model disease progression, and how they may be used to evaluate medical hypotheses related to disease progression pathways and the effect of medical interventions on those pathways.

We further hypothesize that the translation of general features learned by clinical LLMs during pretraining to fine-tuned tasks is directly related to the quality of these clinical latent spaces in modeling phenotypes and temporal locality. Specifically, we propose that implicit organization with respect to "clinically-aware"" designs *improves proportionally with the total number and temporal resolution of patient trajectories observed during pretraining*.

However, significant roadblocks exist in confirming the emergence of trajectories along longitudinal datasets, particularly in scaling to populations past tens of millions of patients. To contextualize the engineering feat required, consider the U.S. Department of Veterans Affairs (VA) which houses one of the largest centralized repositories of clinical data in their Corporate Data Warehouse (CDW). As of 2023, the CDW has archived clinical documents for 4.18 B admissions from nationwide inpatient and outpatient admissions for over 13.8 million patients spanning from January 1, 2000, to January 1, 2022. The urgent need for scalable data analysis is underscored by the VA’s estimated growth rate for their clinical corpus, projected to increase by X00 million documents per year. Even so, the high longitudinal resolution of the VA dataset makes it an important dataset for foundation modeling with an average of 131.9 admissions per patient and over 4.98 M patient years worth of observational data (3 orders of magnitude greater than the 4.47 K patient years covered in MIMIC-IV).

#### Maps Of Disease Progression

As advancements in computational medicine and artificial intelligence continue to unfold, we anticipate the detection of disease progression will be enhanced through insights derived from large-scale longitudinal datasets and applied foundation modeling, ranging from identifying predisposition for chronic conditions to precipitation of acute illnesses. Mapping disease progression would revolutionize public health strategies enabling policymakers and healthcare professionals to implement targeted interventions and preventive measures tailored to the specific stages of disease development. In identifying early indicators and risk factors associated with disease progression, public health initiatives could be optimized to intervene at critical junctures, thereby reducing the burden of illness on nationwide healthcare systems and improving population health outcomes.

Given sufficient healthcare data, these maps may help understand the rapid collapse of health among frail or high comorbid patients^58^. Additionally, maps may help disentangle the complex interplay between genetic predispositions, environmental factors, and medical interventions through infusion with multiple healthcare modalities. Clinicians can gain insights into the dynamics of disease, allowing for early detection, risk stratification, and targeted interventions. Moreover, these maps could serve as invaluable tools for predicting disease trajectories, optimizing treatment strategies, and identifying novel therapeutic targets.

However, with these newfound repositories of clinical knowledge comes ethical considerations and societal responsibilities. It’s imperative to ensure accuracy and reduced bias on downstream finetuning applications using foundation models, equitable access to healthcare resources and interventions which employ these systems, and guards against the misuse of personal health data. As tools for understanding the dynamics of disease, maps of disease progression may iteratively attain capabilities that extend beyond traditional healthcare applications. The ability to predict disease trajectories, identify vulnerable populations, and anticipate the spread of illnesses may hold implications for shaping societal well-being in positive or negative ways. Methods of ascertaining clinical capability as a function model complexity need to be at the forefront of applied foundation modeling in computational medicine.

## Data Availability

All data produced in the present study are available upon reasonable request to the authors

